# Neutralisation hierarchy of SARS-CoV-2 Variants of Concern using standardised, quantitative neutralisation assays reveals a correlation with disease severity; towards deciphering protective antibody thresholds

**DOI:** 10.1101/2021.05.24.21257729

**Authors:** Diego Cantoni, Martin Mayora-Neto, Angalee Nadesalingham, David A. Wells, George W. Carnell, Luis Ohlendorf, Matteo Ferarri, Phil Palmer, Andrew C.Y. Chan, Peter Smith, Emma M. Bentley, Sebastian Einhauser, Ralf Wagner, Mark Page, Gianmarco Raddi, Helen Baxendale, Javier Castillo-Olivares, Jonathan Heeney, Nigel Temperton, on behalf of the HICC Consortium

## Abstract

**Background:** The rise of SARS-CoV-2 variants has made the pursuit to define correlates of protection more troublesome, despite the availability of the World Health Organisation (WHO) International Standard for anti-SARS-CoV-2 Immunoglobulin sera, a key reagent used to standardise laboratory findings into an international unitage.

**Methods:** Using pseudotyped virus, we examine the capacity of convalescent sera, from a well-defined cohort of healthcare workers (HCW) and Patients infected during the first wave from a national critical care centre in the UK to neutralise B.1.1.298, variants of interest (VOI) B.1.617.1 (Kappa), and four VOCs, B.1.1.7 (Alpha), B.1.351 (Beta), P.1 (Gamma) and B.1.617.2 (Delta), including the B.1.617.2 K417N, informally known as Delta Plus. We utilised the WHO International Standard for anti-SARS-CoV-2 Immunoglobulin to report neutralisation antibody levels in International Units per mL.

**Findings:** Our data demonstrate a significant reduction in the ability of first wave convalescent sera to neutralise the VOCs. Patients and HCWs with more severe COVID-19 were found to have higher antibody titres and to neutralise the VOCs more effectively than individuals with milder symptoms. Using an estimated threshold for 50% protection, 54 IU/mL, we found most asymptomatic and mild cases did not produce titres above this threshold.

**Interpretation:** Expressing our data in IU/ml, we provide a benchmark pre-vaccine standardised dataset that compares disease severity with neutralising antibody titres. Our data may now be compared across multiple laboratories. The continued use and aggregation of standardised data will eventually assist in defining correlates of protection.

**Funding:** UKRI and NIHR; grant number G107217

**Research in context:** *Evidence before this study:* During the first wave outbreak, much focus was placed on the role of neutralising antibodies and titres generated upon infection to ancestral SARS-CoV-2. Due to the large amounts of different assays used to elucidate the antibody-mediated immunity and laboratory to laboratory, large amounts of invaluable data could not be directly compared in order to define a correlate of protection, due to variability in the results. The WHO International Standard for anti-SARS-CoV-2 Immunoglobulin sera was made in order to standardise future data so that comparisons may take place.

*Added value of this study:* Our study compares the neutralisation capacity of sera from patients and healthcare workers (HCWs) from the ancestral strain of SARS-CoV-2 against new variants, including the current variants of concern in circulation. We also provide data in International Units per mL, a standardised unitage, for infected individuals that have a clinical severity score, allowing us to assess levels of neutralising antibodies across different severities of COVID-19 disease. By providing a method to calibrate most of the variants of concern so that the WHO International Standard for anti-SARS-CoV-2 Immunoglobulin reagent could be used to standardise our results, therefore making them comparable to other laboratories who also standardised their data in an identical manner.

*Implications of all the available evidence:* Continual use and accumulation of standardised data would eventually lead to defining the correlates of protection against SARS-CoV-2. This could help to inform medical staff to identify which individuals would be a greater risk of a potential reinfection to SARS-CoV-2.

## Introduction

SARS-CoV-2 is the causative agent of the COVID-19 pandemic, resulting in more than 200 million cases and over 4 million deaths (1). Since the start of the outbreak in late 2019, the extensive sequencing of circulating virus has revealed the gradual evolution of variants, emerging independently in many countries around the world. Coronaviruses are enveloped viruses with single-stranded positive-sense RNA genomes ranging from 26 to 32 kilobases in length. SARS-CoV-2 is a member of the β-coronavirus genus which also comprises SARS-CoV (2) and Middle East respiratory syndrome coronavirus (MERS-CoV) (3). As the pandemic progressed, a number of single amino acid mutations in the Spike protein were detected, such as D614G and A222V. The D614G mutation was found to increase the density of Spike protein on virions and increased infectivity (4). The rise of variants in circulation containing several mutations in the viral genome altered several properties of the virus (5). According to several criteria including, increased transmissibility, mortality or morbidity, and the ability to evade natural immunity, these variants have been designated as either Variants of Interest (VOI) or Variants of Concern (VOC). Mutations found in the N-terminus and receptor-binding domain (RBD) of the Spike protein are associated with immune evasion (6– 8). For instance, the E484K mutation in the RBD in several VOCs has been reported to cause up to a ten-fold reduction of neutralisation (9), while the more recent L452R mutation found in B.1.427/B.1.429, a VOC originally detected in California, USA, resulted in a 4 fold reduction (10). Antibodies generated from prior infection or vaccination against the initial virus may provide reduced protection against new variants, giving rise to subsequent waves of infection in regional populations previously impacted by earlier COVID-19 outbreaks (11– 13).

The first notable SARS-CoV-2 variant was linked to an outbreak on a mink farm in Denmark, resulting in a culling program to mitigate risk of spreading (14,15). Referred to as Cluster 5 or B.1.1.298, several different groups of mutations were identified, with the most abundant population containing missense and deletion mutations on the Spike; 69/70del, Y453F and D614G. Shortly after, in September 2020, a new variant was detected in the United Kingdom designated B.1.1.7 (Alpha) which was reported to be more transmissible (16,17). In December 2020, the rise of a new variant designated as B.1.351 (Beta) was detected in South Africa. This new variant has the E484K mutation in the Spike protein that is believed to have a strong impact on antibody evasion (9). A variant designated P.1 (Gamma) was detected in Manaus, Brazil, which also harboured mutations similar to B.1.351, and has been reported to also evade antibodies in previously infected individuals (11,13,18). Most recently, the B.1.617.2 (Delta) variant originating from India has rapidly expanded in many countries (19), becoming the dominant VOC in the United Kingdom (20) and shows reduced neutralisation against vaccination (21,22). There are currently several VOIs that are being monitored by the WHO, including the B.1.617.1 variant (Kappa), with the list constantly being updated. It is of high importance to assess the effectiveness of antibodies from individuals who have recovered from natural infection, as this would allow us to ascertain whether natural infection from the early Wuhan virus isolates, herein referred to as ancestral strain, may offer protection against the newly circulating VOCs, as well as assessing the efficacy of neutralising antibodies generated from vaccines. Having this information would be very informative to develop our understanding of SARS-CoV-2 immune correlates of protection (Figure 1), since neutralising antibody levels are predictive of immune protection (23,24).

**Figure 1.**
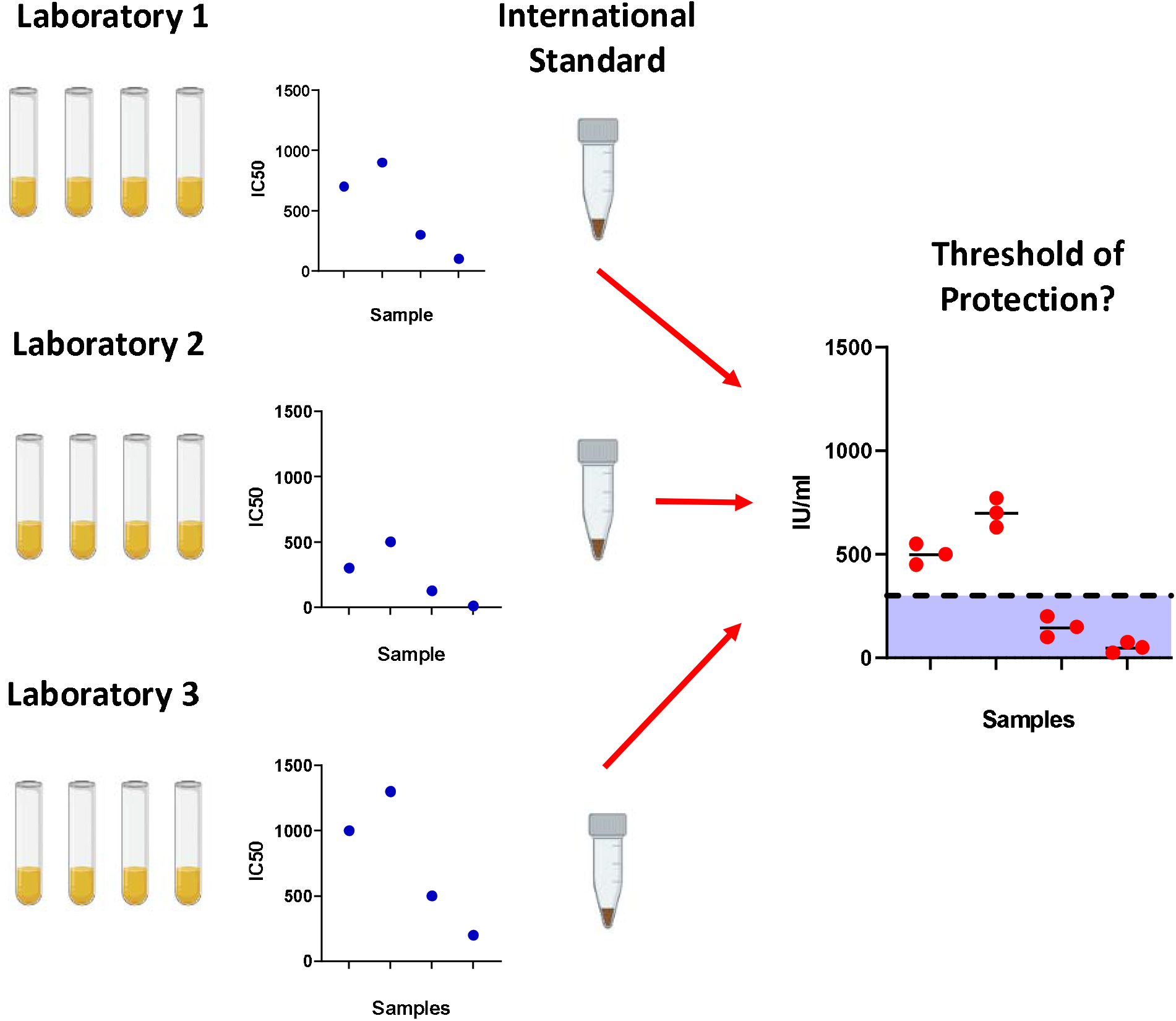
Importance of using the World Health Organisation International Standard serum. To prevent laboratory to laboratory variability between assays, the International Standard was created to standardise results which would allow for cross laboratory comparisons. With gradual accumulation of data, this would permit further analysis into determining correlates for protection against SARS-CoV-2.

Here, we assessed antibodies in sera from convalescent Health Care Workers (HCWs) and patients who were infected during the first wave in the United Kingdom in early 2020. Using well defined and cross validated lentiviral based pseudotyped viruses bearing the ancestral Spike protein from SARS-CoV-2, B.1.1.298, VOI; B.1.617.1 (kappa), and VOCs; B.1.1.7 (Alpha), B.1.351 (Beta), P.1 (Gamma) and B.1.617.2 K (Delta). We also included the B.1.617.2 K417N variant informally named as Delta Plus (Table 1). Pseudotype virus neutralisation assays were performed, reporting IC_50_ values in International Units (IU) according to WHO recommendations (25).

**Table 1.**
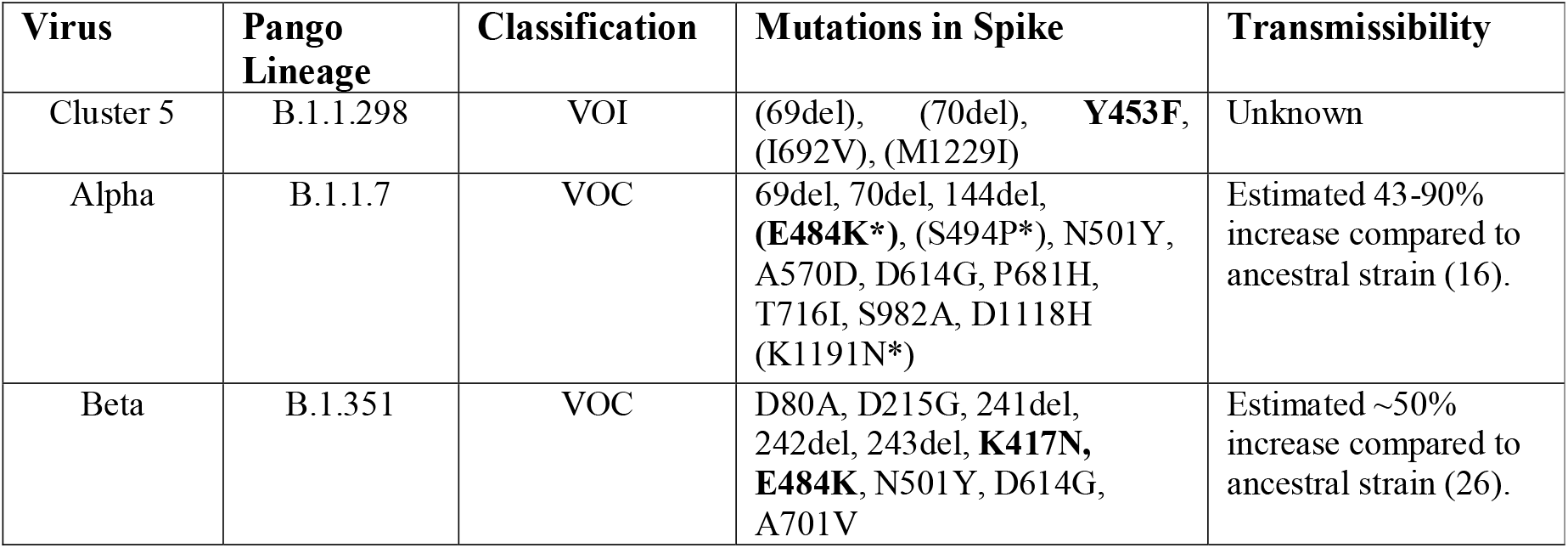
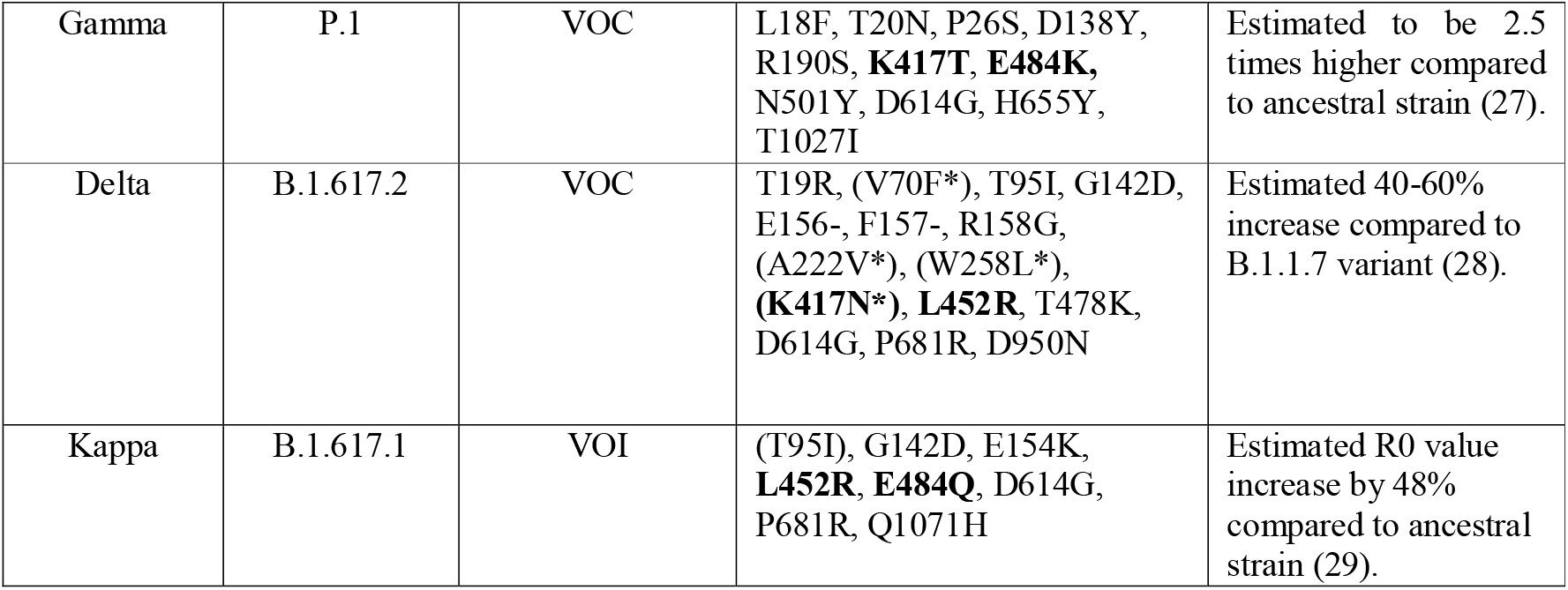
Summary of VOC/VOIs used in this study. Table adapted from the Centre for Disease Control website. Mutations in brackets signify detection in some but not all viral sequences. Delta Plus is an informal name for the delta variant containing the K417N mutation. Plasmids used for this study bearing the specific mutations are listed in the supplementary file. Bold signify mutations implicated in immune escape.

## Materials and Methods

### Tissue Culture

Human Embryonic Kidney 293T/17 (HEK293T17) cells were maintained in DMEM with 10% foetal bovine serum, 1% penicillin/streptomycin and incubated at 37°C and 5% CO_2_.

### Serum Collection

Serum and plasma samples were obtained from HCWs and patients referred to the Royal Papworth Hospital, Cambridge, UK (RPH) for critical care. COVID-19 patients hospitalised during the first wave and as well as NHS healthcare workers working at RPH served as the exposed HCW cohort (Study approved by Research Ethics Committee Wales, IRAS: 96194 12/WA/0148. Amendment 5). NHS HCW participants from the Royal Papworth Hospital were recruited through staff email over the course of two months (20^th^ April 2020-10^th^ June 2020) as part of a prospective study to establish seroprevalence and immune correlates of protective immunity to SARS-CoV-2. Patients were recruited in convalescence either pre-discharge or at the first post-discharge clinical review. All participants provided written, informed consent prior to enrolment in the study. Sera from NHS HCW and patients used in this study were collected between July and September 2020, approximately three months after they were enrolled in the study. Clinical assessment and WHO criteria scoring of severity for both patients and NHS HCW was conducted following the ‘COVID-19 Clinical Management: living guidance (https://www.who.int/publications/i/item/WHO-2019-nCoV-clinical-2021-1). Scoring is based on progression of respiratory disease and cardiovascular collapse: 1=asymptomatic, 2=mild disease, 3=moderate pneumonia, 4 = severe pneumonia, 5=adult respiratory distress syndrome, 6=sepsis, 7= septic shock.

For cross-sectional comparison, representative convalescent serum and plasma samples from seronegative HCWs, seropositive HCWs and convalescent PCR-positive COVID-19 patients. The serological screening used to classify convalescent HCW as positive or negative was done according to the results provided by a CE-validated Luminex assay detecting N-, RBD- and S-specific IgG, (30) a lateral flow diagnostic test (IgG/IgM) and an Electro-chemiluminescence assay (ECLIA) detecting N- and S-specific IgG. Any sample that produced a positive result by any of these assays was classified as positive. The clinical signs of the individuals from which the sample was obtained ranged from 0 to 7 according using the WHO classification described above. Thus, the panel of convalescent serum samples (3 months post-infection) were grouped in three categories: a) Patients (n=38); b) Seropositive Staff (n=24 samples); and c) Seronegative Staff (n=39). Age, sex and symptom severity score is shown in Table 1.

### Generation of Spike expression plasmids

The ancestral strain SARS-CoV-2 Spike expression plasmid (pcDNA3.1+) is based on the Wuhan-Hu-1 sequence and was kindly gifted by Professor Xiao-Ning Xu, Imperial College, London. Mutations of each variant sequence were identified via website databases NexStrain (31), Pango Lineages (32,33) and Centre for Disease Control (CDC) (34). The P.1 variant Spike expression plasmid (pEVAC) was synthesised commercially (GeneArt) with a 19 amino acid C-terminus truncation to increase yields in pseudotyped virus production. The Spike expression plasmids of B.1.1.7 (pI.18), B.1.351 (pI.18) and B.1.1.298 (pcDNA3.1+) were generated in-house by site directed mutagenesis. B.1.617.1 (pcDNA 3.1+) and B.1.617.2 (pcDNA 3.1+) Spike plasmids were kindly donated by Dalan Bailey, Pirbright Institute, G2P Consortium. B.1.617.2 K417N was generated in house by site directed mutagenesis. All plasmids were sequenced to verify successful generation of mutations.

### Pseudotype virus generation

We generated pseudotyped viruses (PVs) bearing the Spike protein of the SARS-CoV-2 Wuhan Type and VOCs as previously described (35). Briefly 1000ng of p8.91 HIV Gag-pol, 1500ng of pCSFLW luciferase and 1000ng of SARS-CoV-2 Spike plasmids were resuspended in Opti-MEM and mixed with FuGENE HD (Promega) at a 1:3 ratio. Transfection complexes were then added dropwise in T-75 culture flasks containing HEK293T/17 cells with replenished fresh DMEM at 70% cell confluency. The culture media was harvested 48 hours post transfection and filtered through a 0.45µm cellulose acetate filters. PVs were then titrated and aliquoted for storage at -70°C.

**Pseudotype virus titration**

The day prior to titration, HEK293T/17 cells were transfected with human ACE-2 (pcDNA 3.1+) and TMPRSS2 (pcDNA 3.1+) expression plasmids using FuGENE HD, to render cells permissible to PVs bearing the SARS-CoV-2 Spike protein. On the day of titration, 100µL of undiluted PV supernatant was serially diluted 2-fold down white F-bottom 96-well plates in 50µL of DMEM. HEK293T/17 cells expressing ACE/TMPRSS2 were added at 10,000 cells per well. Plates were incubated for 48 hours at 37°C and 5% CO_2_. After incubation, the media was aspirated, and cells were lysed using Bright-Glo reagent (Promega) and luminescence was measured using a GloMax luminometer (Promega). PV entry was measured based on relative luminescence units per ml (RLU/ml).

### Neutralisation Assays

Pseudotype microneutralisation assays (pMN) were carried out as previously described (35). Briefly, human convalescent serum was mixed with DMEM at a 1:40 input dilution and serially diluted 2-fold to 1:5,120 in a white F-bottom 96 well plate. PVs were added at a titre around 5×10^6^ RLU/ml in each well. Plates were incubated for one hour at 37°C and 5% CO_2_, followed by addition of HEK293T/17 cells expressing ACE2/TMPRSS2 at 10,000 cells per well. Plates were incubated for 48 hours prior to assaying with Bright-Glo reagent. Each experiment was performed alongside either the NIBSC 20/162 calibrant, HICC-pool 2 and HICC-pool 3, internal calibrants generated from a pool of serum samples from patients. IC_50_ values below 1:40 dilution were considered negative.

### Calculation of International Units from IC_50_ values

IC_50_ values were calculated for the neutralisation assays based on 4-parameter log-logistic regression dose response curves. These curves were fit using AutoPlate (Palmer et al, under review) and the R package drc (36). Before converting IC_50_ values into International Units we demonstrated that the assumption of parallel lines was met for different calibrants against each tested variant. For each variant we fit two models one allowing each calibrant to have its own IC_50_ value and its own gradient and one where a single gradient was shared between calibrants. These two models were compared using an ANOVA test.

After demonstrating parallelism between internal calibrants and the WHO International Standard, we calculated the units of our calibrants. The WHO International Standard (NIBSC code 20/136) has a potency of 1000 IU/ml for neutralising antibody activity after reconstitution. We determined the International Units of our internal calibrants against the ancestral virus and VOCs as a ratio of the calibrant’s potency relative to 20/136.

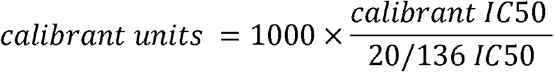

To convert the IC_50_ of samples to International Units, we calculated the sample’s potency as a ratio relative to that determined for the internal calibrant.

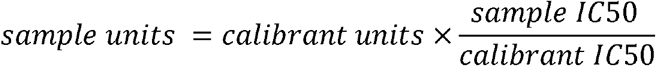

For measurements where the IC_50_ dilution was less than the minimum tested dilution (1:40) the IC_50_ value was set to zero. To avoid these samples dominating calculation, 1 was added to all values when calculating geometric means for IC_50_ dilutions and International Units.

International Units allow neutralisation measured in one laboratory against a specific strain to be compared with that measured in a different laboratory. However, it cannot be used to compare neutralisation between different variants.

### Statistical methods

Dose response curves were fit to pMN data using AutoPlate software (*Palmer et al*, under review). To identify escape mutants, we compared how easily different variants could be neutralised by convalescent sera from patients and previously infected HCWs. Sample potency (IC_50_) was compared between each variant and the ancestral strain using a paired one-sided Wilcoxon signed rank test in R (37,38). Our one-sided test assumed that the ancestral strain was more potently neutralised than the other VOCs.

We compared neutralisation (IU/ml) by our patient and previously infected HCW cohorts of each VOC using an unpaired test, Wilcoxon rank sum test (37,38). We used a one-sided test which assumed that patients would show greater IC_50_ values.

We also tested whether the difference in IC_50_ between patients and previously infected HCWs was the same for different variants. For this we fit a linear mixed model in ‘lme4’ (39) predicting the natural log of the IC_50_ based on cohort and the variant being neutralised. A random intercept was used to account for measuring each sample against five variants. Only measurements with detectible neutralisation were included in this analysis. After filtering out non-neutralising measurements and log transformation, visual investigation of the residuals showed no trends or violations of the assumption of normality. We also fit a second model with an interaction to allow the effect of cohort to differ between variants. The significance of this interaction was assessed by comparing the two models using an F-test based on the Kenward-Rodger correction (40).

Finally, we investigated how disease severity was related to IC_50_ in all variants for samples with detectible neutralisation. For this we used a linear mixed model similar to the one described above but using WHO clinical COVID-19 severity scores on the combined group of HCWs and patients.

## Results

**Table 1.**
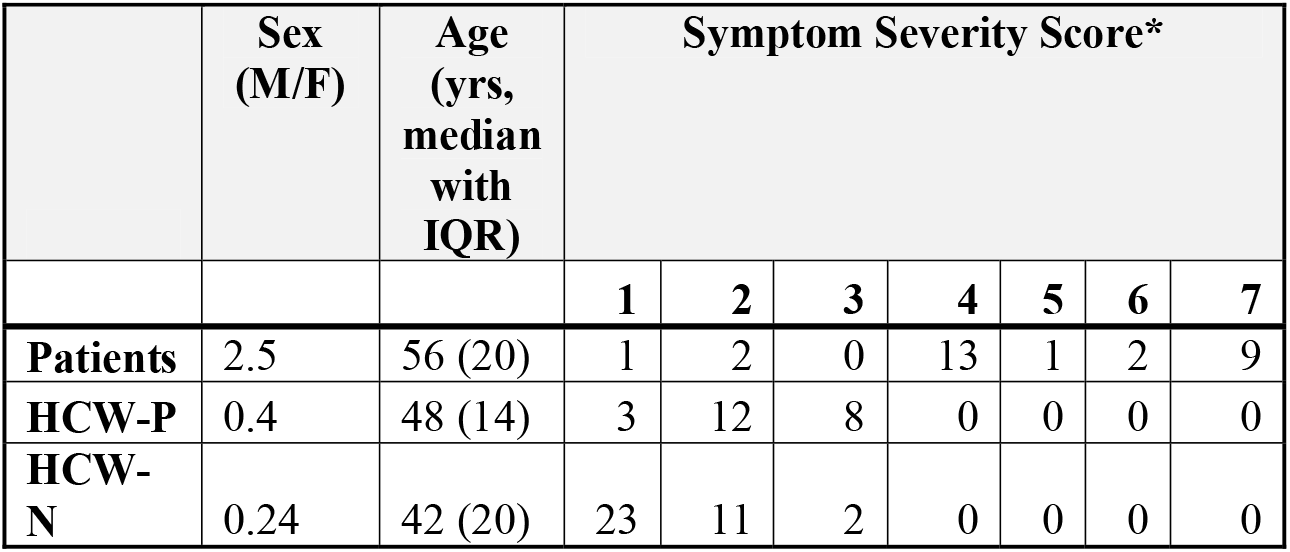
Cohort Demographic and Severity Score Classification. *Symptom severity score: ‘COVID-19 Clinical Management: living guidance (https://www.who.int/publications/i/item/WHO-2019-nCoV-clinical-2021-1).

### Neutralisation Responses to circulating SARS-CoV-2 variants

To assess the neutralisation activity of antibodies in convalescent serum from patients (n=38) and previously infected, seropositive HCWs (n=23), pMN were conducted with PVs bearing the ancestral Spike or VOCs (Figure 2). Compared to the neutralisation titres against PVs bearing the ancestral Spike, we observed the following geometric mean fold changes in neutralisation titres: B.1.1.298: 1.1 fold decrease, B.1.1.7: 1.8 fold decrease, B.1.617.2 K417N: 3.1 fold decrease, B.1.617.2: 4.8 fold decrease, B.1.617.1: 4.9 fold decrease, P.1: 8.2 fold decrease and lastly, B.1.351: 8.3 fold decrease. Our data shows that VOCs P.1 and B.1.351 have the greatest decrease in neutralisation, consistent with previous reports (12). We also report that the VOI B.1.617.1 and B.1.617.2 VOC are similarly neutralised. Lastly, we found that the K417N mutation in the B.1.617.2 Delta Plus increased the neutralisation titres compared to B.1.617.2 delta VOC.

**Figure 2.**
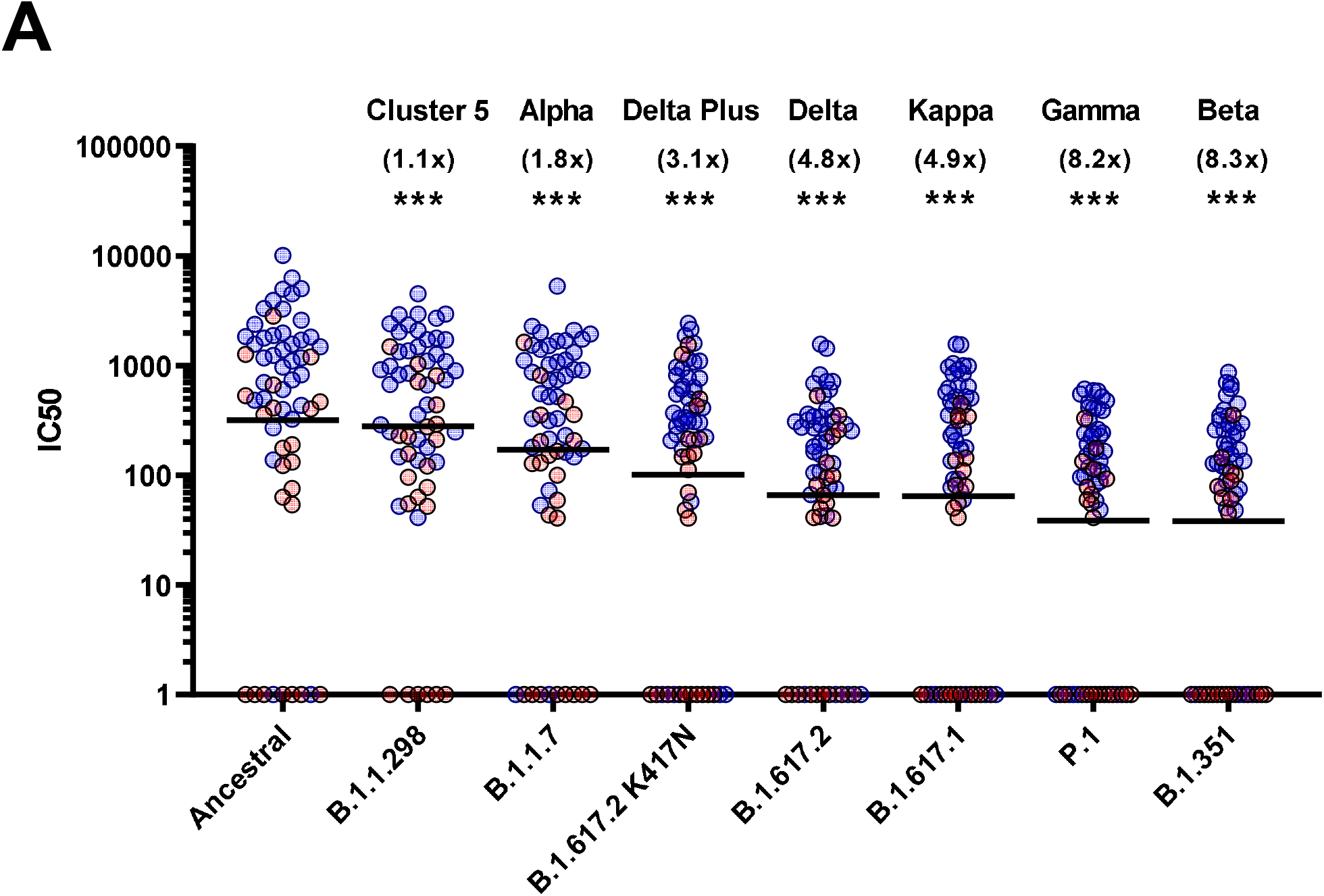
Neutralisation of SARS-CoV-2 pseudotypes by convalescent Serum from seropositive hospital patients and health care workers. Neutralisation assays were carried out using pseudotypes expressing either ancestral spike or B.1.1.298, B.1.1.7, B.1.617.2 K417N, B.1.617.2, B.1.617.1, P.1 and B.1.351 (A). Data is presented in order of increasing fold changes (values in brackets) against the ancestral strain, revealing that VOCs B.1.351 and P.1 have the largest fold decreases (8.2 and 8.3 fold decrease respectively. Wilcoxon signed rank tests were used for statistical analysis between ancestral strain and each VOC (p=<0.001). Black lines denote geometric means. Blue circles represent samples derived from patients, red circles represent samples derived from previously infected healthcare workers.

### Sub-cohort analysis reveals increased antibody neutralisation titres in patients

Sub-cohort analysis was used to evaluate antibody titre between patients and healthcare workers (HCWs). The results reveal that the patients (n=38) had more potent neutralising antibodies against all variants that previously infected HCWs (n=23) (Figure 3) (p<0.001). The geometric mean of IC_50_ values of previously infected HCWs against the ancestral strain was closest to that of non-infected HCWs (n=36) for B.1.351 and P.1 variants. These data suggest that VOC B.1.351 and P.1 are less sensitive to neutralising antibodies found in individuals with a history of asymptomatic infection or mild disease. We used the WHO International Standard to convert all IC_50_ values into IU/ml to allow for inter-laboratory comparison (Table 2). Due to differing immunoreactivities, each of the variants were independently calibrated to the International Standard. VOI B.1.617.1 and VOCs B.1.617.2 and VOC B.1.617.2 K417N IC_50_ values were not calibrated as we were unable to demonstrate parallelism between the curves as described in the methods section.

**Figure 3.**
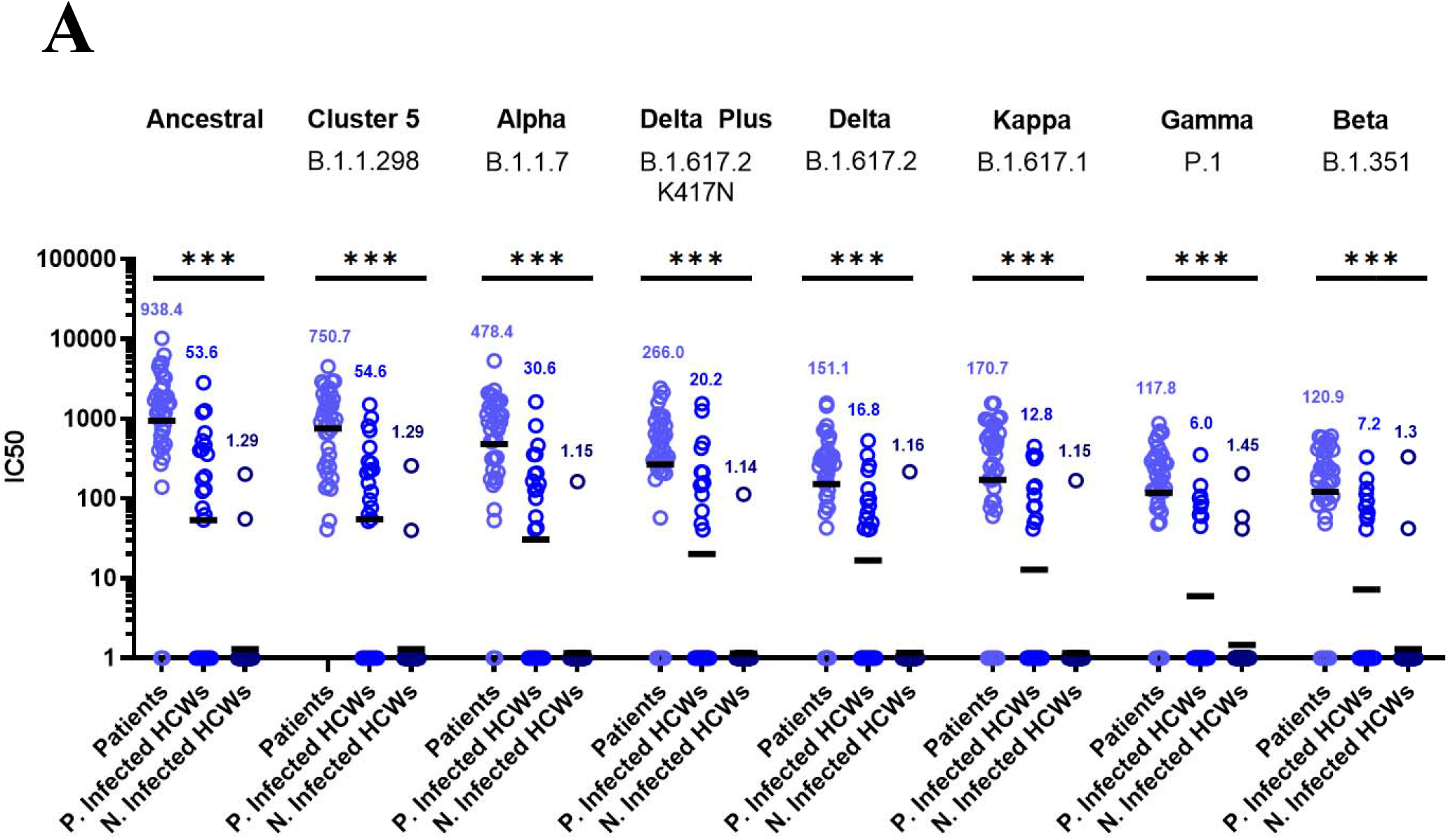
Neutralisation titres split by well defined patient and healthcare worker cohorts. When neutralisation titres were split into cohorts of patients, previously infected HCWs and non-infected HCWs, we observe higher neutralisation titres amongst patients across all variants (A). ANOVA tests were used for statistical analysis between the cohort groups (p=<0.001). Black lines denote geometric means. Geometric means are reported above the datasets.

### Disease severity correlates with antibody neutralisation titres

We wanted to observe the correlations between disease severity of infected individuals and antibody neutralisation titres against the ancestral strain of SARS-CoV-2. The IC_50_ titres were converted into IU/mL and graphed against the clinical COVID-19 severity scores to allow for reproducibility and to compare against an estimated 50% protective threshold defined in literature at 54 IU/mL (95% CI 30-96 IU/mL) (23). Our data shows a clear relationship between disease severity and neutralisation potency against SARS-CoV-2. We also observed 23 samples having neutralising antibody titres below the predicted 50% protective threshold, most of which have a disease severity score from 1 to 3. All samples tested above 4 on the severity score have neutralising antibody titres above the predicted 50% protective threshold.

### Disease severity correlates with higher IC_50_ titre across the VOIs and VOCs

Finally, we tested whether IC_50_ was correlated with the WHO clinical criteria of COVID-19 severity and if this relationship was the same for all VOCs (Figure 4B). CCOVID-19 severity was significantly correlated with IC_50_, although this relationship did not differ between VOCs (severity F(1, 43.2)=18.5, p<0.0001; interaction F(7, 249)=1.29, p=0.26). As before the IC_50_ values were log transformed. Note that this means we tested proportionate, rather than absolute decrease in neutralising IC_50_

**Figure 4.**
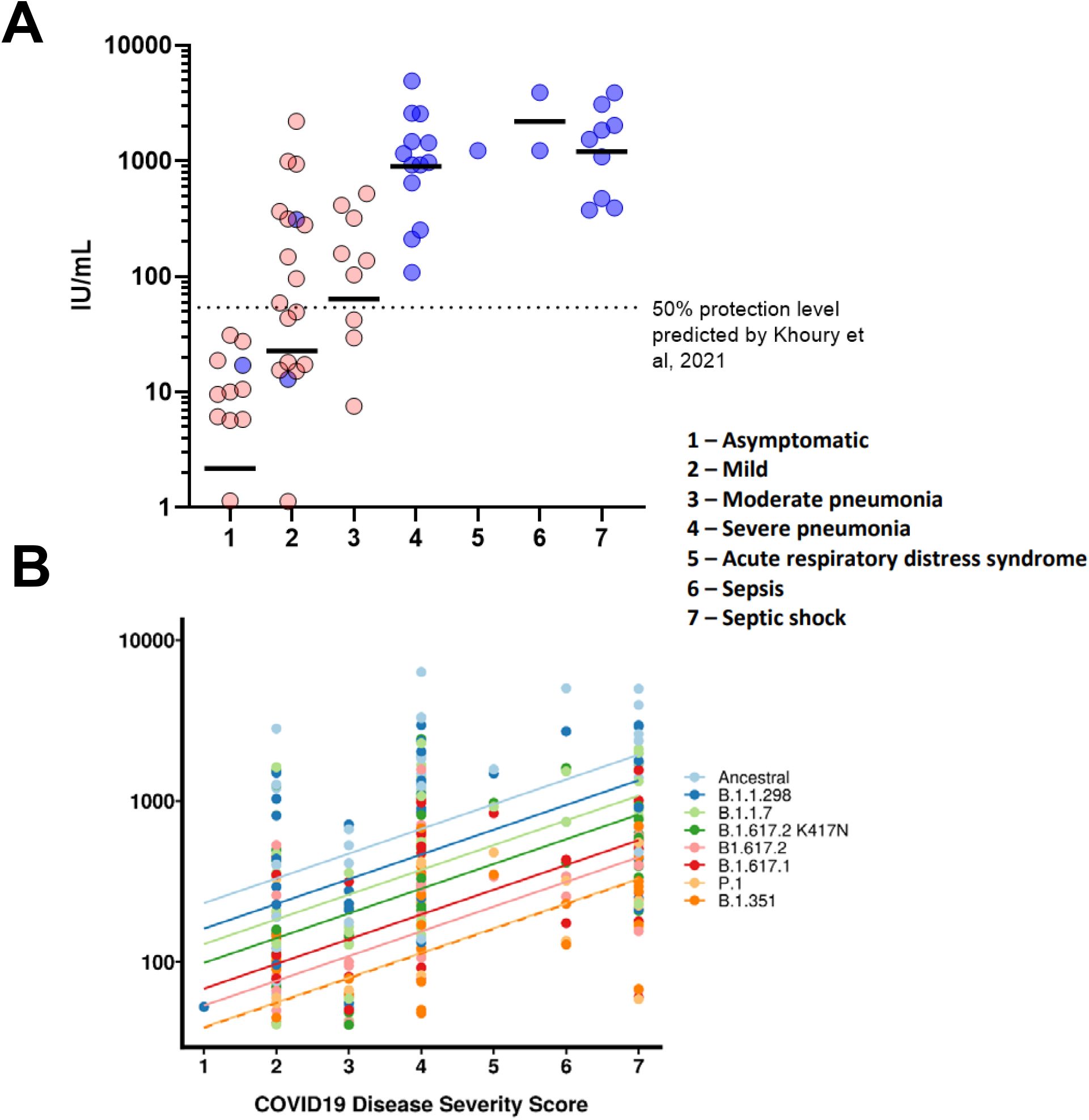
COVID-19 disease severity is associated with increased neutralising antibody titres. IC_50_ titres from patients and HCWs were converted into IU/ml and plotted against the severity of COVID-19 disease using a scoring system. Using pseudoviruses expressing the ancestral spike, we observed a correlation between severity of COVID-19 and neutralisation potency, reaching a plateau at severity scores 4 (severe pneumonia) to 7 (septic shock) (A). Asymptomatic individuals had the lowest titres of nAbs. Blue circles represent samples derived from patients, red circles represent samples derived from previously infected healthcare workers. To compare IC_50_ titres from pseudotypes expressing all VOCs spike, IC_50_ was used as the units of neutralisation as IU/ml does not allow for comparisons against variants (B).

## Discussion

The COVID-19 pandemic has resulted in multiple nationwide lockdowns and renewed efforts to accelerate vaccination programs around the globe. There is a strong urge to return to normality to mitigate further damage to livelihood and economies. Several governments have lowered or dropped COVID-19 restrictions, such as mandatory masks and reopening of bars and restaurants, instead relying on the high vaccination rate of their population to keep cases and hospitalisations low. While there is growing evidence that in the few countries with progressive immunisation programmes there is currently decreasing clinical cases and hospitalisation, relieving pressure on health care infrastructure, there remains a concern that the VOCs may continue to circulate and evolve resistance to vaccine induced immunity. Several reports from Israel found an increased incidence of vaccine breakthrough by the B.1.351 amongst vaccinees (41) and also by B.1.1.7 (42), of which the latter VOC accounted for 94.5% of SARS-CoV-2 isolates in Israel. As a result, understanding the role of pre-existing natural immunity in reducing disease severity is a key factor for informing policies of governments eager to reopen their economies.

The emergence of variants has become a significant issue. One of the first variants, B.1.1.298 also known as the mink variant, contained a unique mutation, Y453F, in the RBD, which was found to enhance ACE2 binding affinity (43). There are several reports that showed a decrease in neutralisation capabilities of antibodies generated by either infection or vaccination with the ancestral wild type against the B.1.1.298 variant, which is consistent with our data. (44,45). Nevertheless, due to the massive culling of mink in farms in Denmark, it is widely believed that the incidence of spillover of this variant from mink to humans has been largely eliminated.

The B.1.1.7 variant was the most prevalent circulating VOC in the United Kingdom, until being recently surpassed by B.1.617.2 (20). Initially, the clinical significance of B.1.1.7 was initially uncertain (45–48). Several studies have investigated whether B.1.1.7 escapes immune evasion from antibodies generated by vaccination (49,50). Overall, most studies have shown a modest decrease in neutralisation from single and double vaccinations against B.1.1.7 (45,46,48,50,51). In our study, the largest reduction in neutralising antibody titres were with P.1 and B.1.351 variants, both of which have a mutational landscape comprising of several amino acids known to affect ACE2 binding and neutralisation (9,12,45,46,52,53). This substantial reduction in neutralisation titres displayed by P.1 and B.1.351 remains to be of concern. This level of immune evasion may lead to susceptibility of reinfection, as has been reported in 3 patients from Brazil with respect to the P.1 variant (11,13,18), and increased likelihood of vaccine breakthrough (41,54). For now, these two VOCs remain to be the most evasive variants known to be in circulation to neutralising antibodies.

The rise of new B.1.617 lineage has resulted in the detection of several sublineages; B.1.617.1, B.1.617.2 and B.1.617.3, of which B.1.617.2 has become the dominant variant in circulation. More recently the K417N mutation was detected in B.1.617.2 in several sequences from Nepal. Our results show that convalescent sera from first wave infected individuals were able to neutralise, albeit with reduce titres, B.1.617.1, B.1.617.2 and B.1.617.2 K417N. In addition, the K417N mutation appears to increase the neutralisation efficiency compared to the original B.1.617.2. The degree of protection against infection to these variants by antibodies derived from ancestral SARS-CoV-2 infection is difficult to gauge, as the neutralising titres were closer to that of B.1.351 or P.1, known to have had reinfection cases, compared to B.1.1.7. Furthermore, several studies have reported similar reductions in neutralisation against variants from the B.1.617 lineages (21,22,55–57). One study has reported significant numbers of vaccine breakthroughs by B.1.617.2 in fully vaccinated HCW in three different hospitals in Dehli, India (22). The authors also observed an increased viral load in these cases, highlighting the fact that the rapid replication rate of B.1.617.2 variant may contribute to vaccine breakthrough by overwhelming an already established immune response.

Standardisation in the reporting of data is critical for comparison of data in different populations and countries and to harmonise assay to assay and lab to lab variability, which will be vital in informing national and international public health policies around the world (58). Here we report our findings in International Units through the use of the WHO International Standard for anti-SARS-CoV-2 Immunoglobulin (NIBSC code: 20/136) comprising a pool of 11 convalescent plasma sourced from the first wave of global infections, when the circulating SARS-CoV-2 sequences were relatively genetically homogeneous. One of the main questions regarding the antiviral neutralising antibody responses remains: what are the immune correlates of protection against SARS-CoV-2 infection? One study has estimated a neutralisation level of approximately 54 IU/mL based on vaccinated populations which was denoted in figure 4A (23). Whilst clinical severity of COVID-19 has already been correlated with neutralising antibody titres (59–62), a lack of a standardised unitage for neutralisation titres means that it is not possible to compare the datasets with current or future correlate of protection predictions. Using the estimate provided by Khoury et al (23), our data shows that asymptomatic and several mild cases from first wave infections are below the estimated 50% level of protection. This does not necessarily mean that these individuals would be reinfected, but rather that their risk of reinfection may be more elevated.

There are several limitations and caveats that are important to mention. Most notably, Khoury et al clearly stated that the 54 IU/mL estimate, the first of its kind for SARS-CoV-2, was based on aggregation of datasets using diverse neutralisation assays and vaccine clinical trial designs which did not use calibrated assays, and asserted that future standardisation was key to defining correlates of protection (23,63). Whilst studies have analysed the degree of correlation between different assays, it is difficult to account for inter-laboratory variation (64). Furthermore, our study only examines a single component of the immune response, and several other markers can be used as correlates of protection such as T cell or B cell responses, of which currently there is no estimated nor defined unitage that correlates with protection. Lastly, a limitation of the WHO International Standard is that it cannot be used to compare data derived from neutralisation assays against different variants due to their individual calibration to the International Unit, based on differing immunoreactivities of the viruses. Whilst calibration can be carried out for variants, assuming parallelism is met during calibration of the curves, the data would be considered standardised and remains comparable to data generated from other laboratories against the same variant. For these reasons, our neutralisation data is kept in IC_50_ when comparisons between variants were made.

In summary, this data, expressed in IU/ml, represents a benchmark “pre-vaccine” standardised dataset comparing infected individuals with different disease outcomes. This will allow multiple laboratories to compare neutralisation potencies calibrated against the WHO International Standard for each studied variant. The continual use of the Standard by various laboratories could greatly increase our ability to establish benchmarks, or thresholds of correlates of immunity against different variants. The next steps involve expanding this standardised data to immunised individuals for comparison of neutralising antibodies in convalescent, versus infected and vaccinated individuals against the different VOCs and establishing thresholds of protection against circulating variants to inform national and international vaccine programmes.

## Data Availability

Raw data will made available upon request.

## Author Contributions

DC, HB, JCO, JH, MP, RW and NT designed the study. DC and MMN carried out the experiments. GR and HB co-ordinated patient and HCW selection, for blood sample collection from RPH. AN, ACYC and PS contributed by processing blood sample into sera, cataloguing and providing reagents. EMB, RW and SE contributed by generating plasmids of VOCs. DAW and PP contributed by processing data and performing statistical analysis of the results. GWC, LO, MF, SE contributed to sample preparation, optimisation of pseudotype virus production, protocol design and interpretation of results. DC wrote the original draft of the manuscript. MMN, AN, DAW, GWC, LO, MF, PP, ACYC, PS, EMB, SE, RW, MP, GR, HB, JCO, JH and NNT provided critical feedback on the data presentation, data analysis and manuscript. All authors had full access to the data generated in this study.

## Data Sharing

Data generated in this manuscript can be made available upon request by contacting the co-corresponding authors.

## Declaration of Interest

We declare no competing interests

## Acknowledgements

This study was undertaken by the Humoral Immune Correlates to COVID-19 (HICC) consortium, funded by the UKRI and NIHR; grant number G107217 (COV0170 - HICC: Humoral Immune Correlates for COVID19). RW and SE received funding from the StMWK (ForCOVID, Bavaria, Germany). We thank the RPH Foundation Trust COVID-19 Research and Clinical teams for supporting recruitment to this study, HCWs and Outpatients who participated in this study. We would like to thank Dr Dalan Bailey at the G2P consortium for providing us with the B.1.617.1 and B.1.617.2 plasmids.

---

**Table 1.** Sub-Cohort Geometric Means and Interquartile Ranges expressed in International Units (IU/ml)

| **Ancestral** | **Patients** | **Previously infected HCWs** | **Non-infected HCWs** |
| --- | --- | --- | --- |
| Geometric Mean | 744.5 | 45.22 | 1.278 |
| Interquartile Range | 1446.9 | 366 | 0 |
| **B.1.1.298 (Cluster 5)** | **Patients** | **Previously infected HCWs** | **Non-infected HCWs** |
| Geometric Mean | 225.2 | 22.42 | 1.209 |
| Interquartile Range | 467.94 | 86.83 | 0 |
| **B.1.1.7 (Alpha)** | **Patients** | **Previously infected HCWs** | **Non-infected HCWs** |
| Geometric Mean | 478.4 | 30.59 | 1.152 |
| Interquartile Range | 1299 | 207 | 0 |
| **B.1.351 (Beta)** | **Patients** | **Previously infected HCWs** | **Non-infected HCWs** |
| Geometric Mean | 503 | 13.17 | 1.672 |
| Interquartile Range | 1209.3 | 398.1 | 0 |
| **P.1 (Gamma)** | **Patients** | **Previously infected HCWs** | **Non-infected HCWs** |
| Geometric Mean | 211.3 | 10.2 | 1.362 |
| Interquartile Range | 691.3 | 170.8 | 0 |

**Table 2.** Sub-Cohort Geometric Means and Interquartile Ranges expressed in International Units (IU/ml) The geometric means and interquartile ranges obtained from the datasets presented in Figure 3 were converted into International Units (IU/ml) to allow for cross laboratory comparisons. The IU/ml cannot be used to cross compare between variants.

|  | COVID-19 Disease Severity Scores |  |  |  |  |  |  |
| --- | --- | --- | --- | --- | --- | --- | --- |
|  | 1 | 2 | 3 | 4 | 5 | 6 | 7 |
| Geometric Mean | 1 | 13.91 | 38.22 | 900.7 | 1230 | 2199 | 1206 |
| Interquartile Range | 0 | 295.5 | 343.9 | 1574.3 |  |  | 2133.2 |

**Table 3.** COVID-19 disease severity scores geometric means and interquartile ranges expressed in International Units (IU/ml) The geometric means and interquartile ranges obtained from the datasets presented in Figure 4A were converted into International Units (IU/ml) to allow for cross laboratory comparisons. Interquartile ranges for datasets in disease severity scores 5 and 6 do not have an interquartile range due to lack of data points.

## Notes

### Competing Interest Statement

The authors have declared no competing interest.

### Clinical Trial

96194 12/WA/0148. Amendment 5

### Author Declarations

Study approved by Research Ethics Committee Wales, IRAS: 96194 12/WA/0148. Amendment 5

