## Supplementary Figure 1 for "Neutralisation hierarchy of SARS-CoV-2 Variants of Concern using standardised, quantitative neutralisation assays reveals a correlation with disease severity; towards deciphering protective antibody thresholds"

**Supplementary File**

| **Variant** | **Mutations** |
| --- | --- |
| B.1.1.298 (Cluster 5) | 69/70del, Y453F, D614G |
| B.1.1.7 (Alpha) | Δ69-70, Δ144, N501Y, A570D, D614G, P681H, T716I, S982A, D1118H |
| B.1.351 (Beta) | D80A, D215G, Δ242-244, K417N, E484K, N501Y, D614G, A701V |
| P.1 (Gamma)* | L18F, T20N, P26S, D138Y, R190S, K417T, E484K, N501Y, D614G, H655Y, F1176V |
| B.1.617.2 (Delta)* | T95I, G142D, E154K, L452R, E484Q, D614G, P681R, Q1071H |
| B.1.617.2 K417N (Delta Plus)* | T95I, G142D, E154K, K417N, L452R, E484Q, D614G, P681R, Q1071H |
| B.1.617.1 (Kappa)* | T95I, G142D, E154K, L452R, E484Q, D614G, P681R, Q1071H |

### Variant mutations

Table S1 description of variant mutations

*contain 19aa C-terminus truncation to increase pseudotype production titres

### Parallel lines analysis

Before calculating the potency of our internal calibrants in international units we had to validate the assumption of parallelism between the dose response curves of our calibrants and the International Standard 20/136. We ensured parallelism for all calibrants against all variants. For each variant we fit two models one allowing each calibrant to have its own IC_50_ value and its own gradient and one where a single gradient was shared between calibrants. These two models were compared using an ANOVA test.

# B.1

Table S2.1 Results of ANOVA comparing model with a single gradient for calibrants (1^st^ model) and allowing separate gradients (2^nd^ model). Non-significant results indicate parallelism between curves against the B.1 variant.

|  | ModelDf | RSS | Df | F value | p value |
| --- | --- | --- | --- | --- | --- |
| 1st model | 57 | 3228.872 | NA | NA | NA |
| 2nd model | 54 | 3130.500 | 3 | 0.565629 | 0.6400322 |

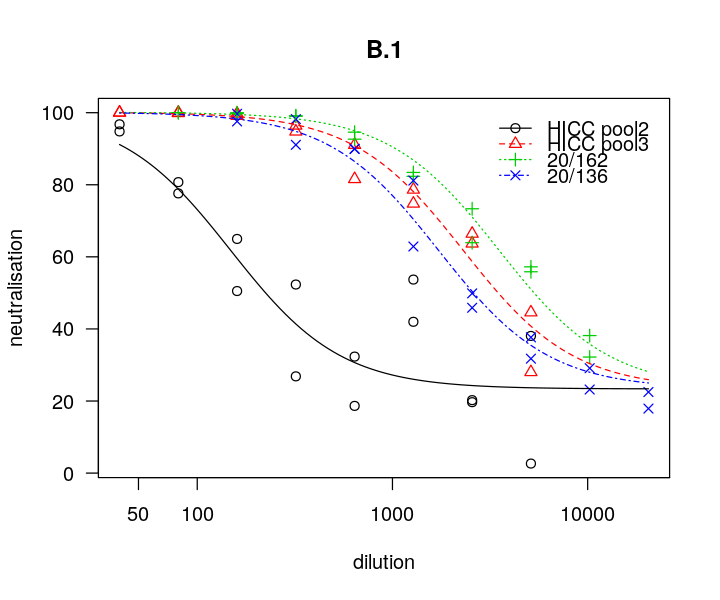
 Figure S1.1 Dose responses curves for three internal calibrants and the International standard against the B.1 variant of SARS-CoV-2.

# B.1.1.298

Table S2.2 Results of ANOVA comparing model with a single gradient for calibrants (1^st^ model) and allowing separate gradients (2^nd^ model). Non-significant results indicate parallelism between curves against the B.1.298 variant.

|  | ModelDf | RSS | Df | F value | p value |
| --- | --- | --- | --- | --- | --- |
| 1st model | 57 | 11813.88 | NA | NA | NA |
| 2nd model | 54 | 10607.99 | 3 | 2.046186 | 0.1182651 |

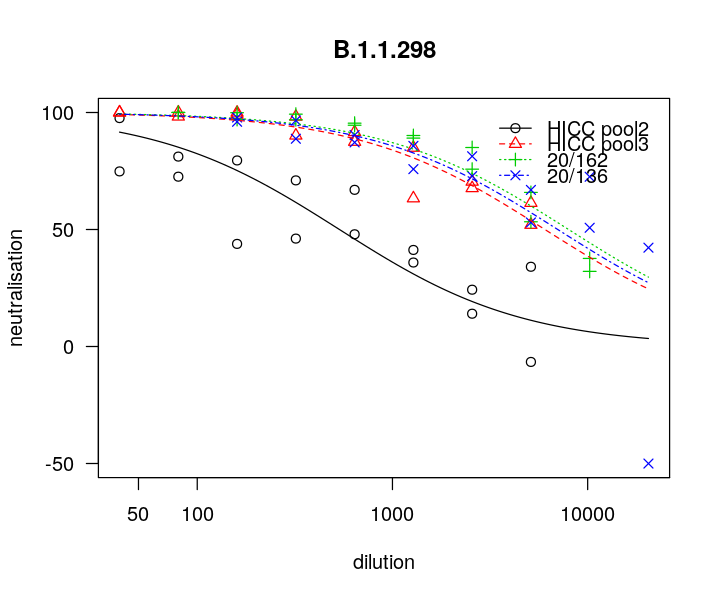
 Figure S1.2 Dose responses curves for three internal calibrants and the International standard against the B.1.1.298 variant of SARS-CoV-2.

# B.1.1.7

Table S2.3 Results of ANOVA comparing model with a single gradient for calibrants (1^st^ model) and allowing separate gradients (2^nd^ model). Non-significant results indicate parallelism between curves against the B.1.1.7 variant.

|  | ModelDf | RSS | Df | F value | p value |
| --- | --- | --- | --- | --- | --- |
| 1st model | 57 | 5378.322 | NA | NA | NA |
| 2nd model | 54 | 4986.303 | 3 | 1.415143 | 0.248 |

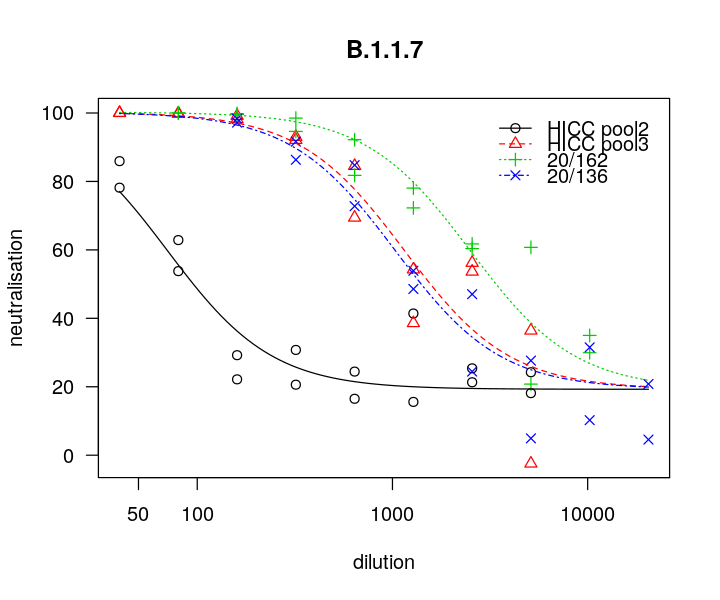
 Figure S1.3 Dose responses curves for three internal calibrants and the International standard against the B.1.1.7 variant of SARS-CoV-2.

# B.1.351

HICC pool 2 showed very low neutralisation against the B.1.351 variant. As such this calibrant was not used to calculate the International Units of samples against the B.1.351 variant.

Table S2.4 Results of ANOVA comparing model with a single gradient for calibrants (1^st^ model) and allowing separate gradients (2^nd^ model). Non-significant results indicate parallelism between curves against the B.1.351 variant.

|  | ModelDf | RSS | Df | F value | p value |
| --- | --- | --- | --- | --- | --- |
| 1st model | 57 | 9643.841 | NA | NA | NA |
| 2nd model | 54 | 8546.210 | 3 | 2.311829 | 0.0863982 |

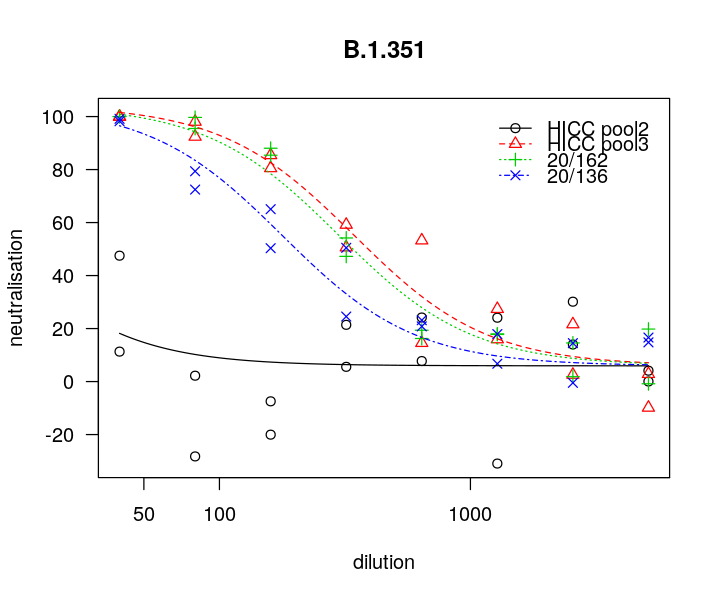
 Figure S1.4 Dose responses curves for three internal calibrants and the International standard against the B.1.351 variant of SARS-CoV-2.

# P.1

Table S2.5 Results of ANOVA comparing model with a single gradient for calibrants (1^st^ model) and allowing separate gradients (2^nd^ model). Non-significant results indicate parallelism between curves against the P.1 variant.

|  | ModelDf | RSS | Df | F value | p value |
| --- | --- | --- | --- | --- | --- |
| 1st model | 57 | 20529.94 | NA | NA | NA |
| 2nd model | 54 | 20289.72 | 3 | 0.213111 | 0.8868899 |

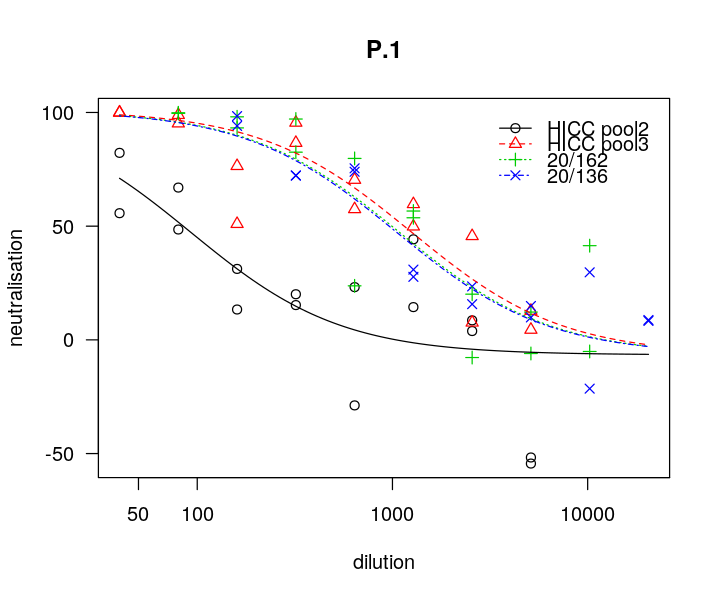

Figure S1.5 Dose responses curves for three internal calibrants and the International standard against the P.1 variant of SARS-CoV-2.

## B.1.617.1

Table S2.6 Results of ANOVA comparing model with a single gradient for calibrants (1^st^ model) and allowing separate gradients (2^nd^ model). Non-significant results indicate parallelism between curves against the B.1.617.1 variant.

|  | ModelDf | RSS | Df | F value | p value |
| --- | --- | --- | --- | --- | --- |
| 1st model | 27 | 2334.280 | NA | NA | NA |
| 2nd model | 26 | 1323.966 | 1 | 19.8405 | 0.0001419239 |

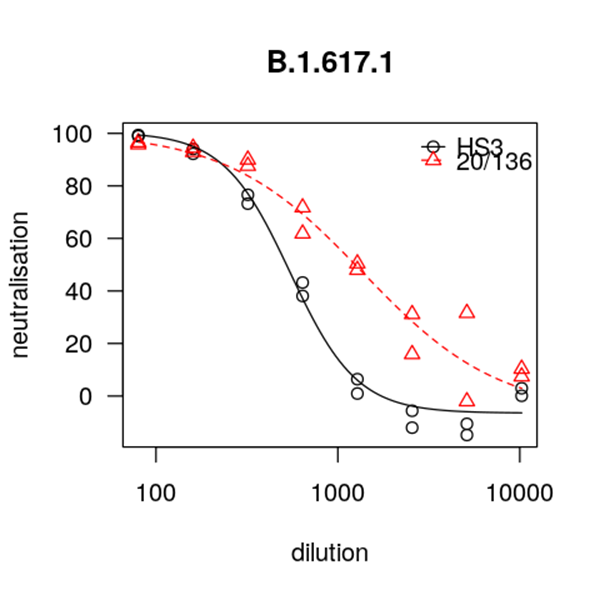

Figure S1.6 Dose responses curves for the International standard against the B.1.617.1 variant of SARS-CoV-2.

## B.1.617.2

Table S2.7 Results of ANOVA comparing model with a single gradient for calibrants (1^st^ model) and allowing separate gradients (2^nd^ model). Non-significant results indicate parallelism between curves against the B.1.617.2 variant.

|  | ModelDf | RSS | Df | F value | p value |
| --- | --- | --- | --- | --- | --- |
| 1st model | 27 | 2934.560 | NA | NA | NA |
| 2nd model | 26 | 2424.049 | 1 | 5.475665 | 0.02723315 |

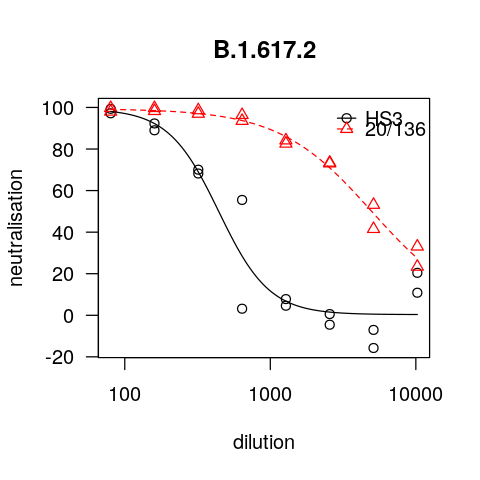

Figure S1.7 Dose responses curves for the International standard against the B.1.617.2 variant of SARS-CoV-2.

## B.1.617.2 K417N

Table S2.8 Results of ANOVA comparing model with a single gradient for calibrants (1^st^ model) and allowing separate gradients (2^nd^ model). Non-significant results indicate parallelism between curves against the B.1.617.2 K417N variant.

|  | ModelDf | RSS | Df | F value | p value |
| --- | --- | --- | --- | --- | --- |
| 1st model | 27 | 9143.489 | NA | NA | NA |
| 2nd model | 26 | 5375.260 | 1 | 18.22683 | 0.0002309578 |

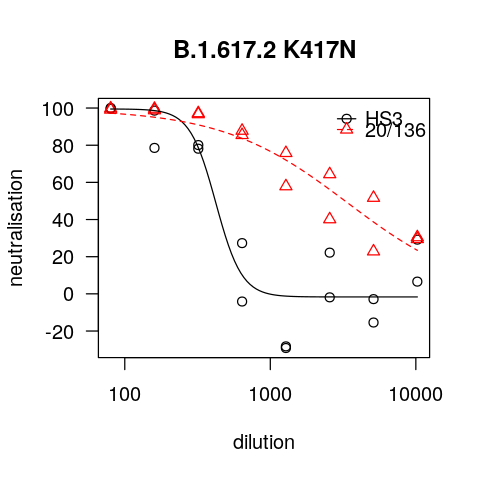

Figure S1.8 Dose responses curves for the International standard against the B.1.617.2 K417N variant of SARS-CoV-2.
